# The impact of cardiorespiratory fitness on Alzheimer’s disease biomarkers and their relationships with cognitive decline

**DOI:** 10.1101/2025.03.03.25323245

**Authors:** AJ Paulsen, I Driscoll, BM Breidenbach, MP Glittenberg, SR Lose, Y Ma, MA Sager, CM Carlsson, CL Gallagher, BP Hermann, K Blennow, H Zetterberg, S Asthana, SC Johnson, TJ Betthauser, BT Christian, DB Cook, OC Okonkwo

## Abstract

**INTRODUCTION:** Relationships between core Alzheimer’s disease (AD) biomarker accumulation and cognitive decline are well-established and the literature generally suggests a favorable relationship of cardiorespiratory fitness (CRF) on AD biomarker accumulation and cognition. Differences in risk of biomarker status conversion or accumulation rates by CRF, or their potential interactive relationships with cognitive decline remain largely unknown.

**METHODS:** Participants (N=533; MeanAGE=65, 70% female) from the Wisconsin Alzheimer’s Disease Research Center and the Wisconsin Registry for Alzheimer’s Prevention underwent serial blood draws, and cognitive and imaging assessments (MeanFollow-up=6.0 years). PET imaging of amyloid-β (Aβ) and tau (T) and plasma phosphorylated tau-217 (pTau-217) were used to determine biomarker status (+/-). Sex-specific estimated CRF (eCRF) tertiles were created using a validated equation. Kaplan-Meier survival curves and Cox-proportional hazards models characterized the risk of becoming biomarker-positive. Linear mixed effects models estimated associations between baseline eCRF and core AD biomarker accumulation and whether eCRF modified relationships between biomarker accumulation and cognitive decline. Analyses were stratified by biomarker +/- status.

**RESULTS:** No significant relationships were observed between eCRF and biomarker trajectories. However, those in the high eCRF group who were also Aβ-(HR[95%CI]=0.42[0.20, 0.88]) and pTau-217-(HR[95%CI]=0.45[0.21, 0.97]) at baseline had a significantly lower risk of becoming biomarker-positive. There was a significant attenuation of the detrimental relationship between Aβ accumulation and cognitive decline for those with high eCRF and Aβ+/T+.

**DISCUSSION:** While CRF did not influence core AD biomarker accumulation trajectories, high CRF did seem to protect against becoming biomarker-positive and attenuate the known deleterious relationship between biomarker accumulation and cognitive decline in Aβ+/T+.

## 1. Introduction

Accumulation of amyloid-β (Aβ) and tau (T), neuropathologic hallmark proteins of Alzheimer’s disease (AD), begins decades prior to clinically relevant symptoms, making them the earliest known markers of AD-related pathological changes.[1–5] Phosphorylated tau (pTau) has also recently been suggested as a key biomarker for the diagnosis of AD, even in preclinical stages.[1,2,6,7] The timing of these pathologic changes is helpful in identifying people at greatest risk while in the preclinical stages along the AD dementia continuum and are highly relevant for research aimed at identifying preventative strategies for AD.[2,3,8]

Age, genetics, and family history of AD are the greatest known risk factors for AD.[9] Conversely, existing literature suggests that higher levels of physical activity (PA) and better cardiorespiratory fitness (CRF) may positively alter both AD neuropathological progression[10–13] and age-related cognitive decline[10,13–17], thereby reducing risk for AD and related dementias. AD biomarker profiles of highly fit individuals seem to be less adversely impacted by genetic risk compared to those less fit.[12] Moreover, the literature suggests that the negative effects of Aβ accumulation on cognitive function are mitigated by higher CRF, and conversely, that downstream effects of physical inactivity moderate the effect of Aβ on cognitive decline.[15,16] The majority of this literature is cross-sectional in nature and limited by modest sample sizes, leaving a knowledge gap in our understanding of the impact of CRF on AD biomarker accumulation. Moreover, trajectories of AD biomarker accumulation differ based on biomarker positivity status (i.e., participants who are already Aβ+ tend to accumulate Aβ at a faster rate than those who are Aβ-, who may not accumulate Aβ at all).[18,19]

The present study examines the relationship between baseline CRF with trajectories of core AD biomarker accumulation, namely Aβ and tau measured using positron emission tomography (PET), and pTau-217 measured in plasma, and the risk of conversion to biomarker positivity for those who were negative at baseline. Additionally, we investigate the potential modification of the relationships between accumulation trajectories of each of these core AD biomarkers and cognitive decline by CRF. Importantly, the present study examines the aforementioned relationships based on AD biomarker status (+/-).

## 2. Methods

### 2.1 Participants

The present sample included 533 participants from the Wisconsin Registry for Alzheimer’s Prevention (WRAP) and the Wisconsin Alzheimer’s Disease Research Center (WADRC), two prospective cohorts enriched for parental history of AD at enrollment, consisting of more than 2000 middle-aged and older adults. Detailed descriptions of enrollment criteria have been previously published.[20,21] The current sample was comprised of participants with available data of interest, predominantly white (92%; 5% Black/African American; 3% all other groups), on average college educated, and cognitively unimpaired at baseline. Participants ranged in age between 46-89 years, with a mean follow-up (SD) of 6.0 (3.9) years (number of visits ranging from 1 to 7). All participants provided written informed consent prior to participation and all procedures were approved by the University of Wisconsin Health Sciences Institutional Review Board.

### 2.2 Cardiorespiratory fitness

An estimated CRF (eCRF) was calculated using a validated equation,[22] which accounts for age, sex, body mass index, resting heart rate, and self-reported PA:

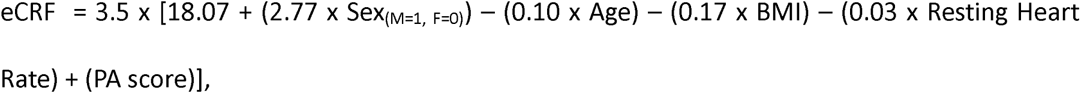

with a resulting estimate of VO_2peak_ in mL/kg/min. BMI was calculated as weight in kilograms divided by height in meters squared. Resting heart rate was measured using a GE Dinamap Pro 400 V2 Vital Signs Monitor (GE Medical Systems Information Technologies, Inc., Milwaukee, WI). Responses to questionnaires characterizing participants’ habitual activity levels (duration, intensity, and frequency per week) were used to calculate the PA score as previously described.[22–24] Briefly, the PA score incorporates the characteristics of habitual PA to assign five distinct PA levels with scores assigned to each level ranging from 0 to 3.03. In addition to the continuous measure of eCRF, a categorical eCRF variable was created by defining sex-specific tertile cut-points across all participants in WRAP/WADRC, which were then applied to the current sample to create low, middle, and high eCRF groups.

### 2.3 Biomarker Measurements

Aβ burden was measured using ^11^C-Pittsburgh Compound B (PiB) PET imaging. PiB acquisition and postprocessing protocols have been previously detailed.[25] Briefly, participants underwent PiB imaging with a 6-minute transmission scan after bolus injection and a 70-minute dynamic scan. Cortical Aβ burden was quantified based on derived distribution volume ratio (DVR) maps using the Logan method with cerebellar gray matter as reference.[25] Amyloid positivity (Aβ+) was defined by a global cortical DVR>1.19.[26] Tau was measured using ^18^F-MK-6240 PET tracer. Acquisition and postprocessing has been previously described.[4] Briefly, standard uptake value ratios (SUVR) were calculated following a 20-minute dynamic acquisition scan. Tau positivity (T+) was defined by tracer uptake in the entorhinal cortex where neurofibrillary tangles (NFT) are expected to first occur (entorhinal SUVR>1.27) or regional tau positivity in NFT associated regions corresponding to Braak staging.[5,27,28]

Participants provided blood samples at each study visit from which pTau-217, which is indicative of Aβ positivity and may also reflect the entire AD pathological process,[29–31] was assayed using ALZpath (ALZpathDX, Carlsbad, CA) at the Clinical Neurochemistry Laboratory in the University of Gothenburg, Sweden. Detailed information on the collection and processing of samples and quantification of pTau-217 has been previously published.[30,31] A binary pTau-217 +/- status was created using a previously determined center-wide reference cut-point of >0.63 pg/mL.[29]

Analyses including pTau-217 (N=445) and tau-PET (N=457) had slightly smaller sample sizes due to missingness of those measures.

### 2.4 Neuropsychological Assessment

Participants undergo biannual neuropsychological assessment as part of their WRAP/WADRC participation. The present study employed a modified Preclinical Alzheimer’s Cognitive Composite (PACC-3) score, comprised of the Rey Auditory Verbal Learning Test Delayed Recall (RAVLT), the Wechsler Memory Scale Logical Memory II, and the Trail Making Test Part B (TMTB), as the utility of global composite scores is superior to performance on individual tests when assessing longitudinal cognitive decline.[32,33] The composite score was z-transformed across the entire WRAP and WADRC cohort for analyses purposes, so that scores among a cognitively unimpaired sample had an approximately normal distribution with mean=0 and SD=1.

### 2.5 Statistical Analyses

All analyses were conducted using RStudio version 2023.12.1+402 (Posit Software, PBC). Analyses were stratified by Aβ-PET status (Aβ- or Aβ+) at baseline given that biomarker accumulation trajectories tend to differ based on baseline Aβ positivity status, and Aβ is considered the earliest indication of AD pathological change.[18,19,27,34] Differences in baseline characteristics by Aβ+/- status were determined using t-tests for continuous variables and chi-squared tests for categorical variable. Analyses of change in cognition were conducted stratified by Aβ and tau+/- status at any timepoint (i.e., Aβ-T-were never positive and Aβ+T+ were positive for both biomarkers at their most recent visit) as the combined pathology confers the greatest risk of cognitive change[27,35,36] and few participants were T+ at baseline in the current sample, limiting power for stratified analyses. The resultant groups were Aβ-T-, Aβ+T-, Aβ-T+, and Aβ+T+. Statistical significance was determined with a two-tailed *p*<0.05.

Linear mixed effects (LME) models were constructed by including baseline eCRF, years of follow-up for biomarker measurement, and their interaction to investigate whether rates of biomarker accumulation over time differ based on eCRF. This relationship was tested using continuous eCRF (per SD increase) and by eCRF group with the low fitness group as the reference. All models were adjusted for baseline age, sex, years of education, *APOE* ε4 carrier status, parental history of AD, years of follow-up, and random intercept and slope per participant.

To evaluate the relationship between eCRF and conversion to biomarker positivity during follow-up for those who were biomarker negative at baseline, Kaplan-Meier survival curves were constructed by eCRF group. Additional Cox-Proportional Hazards models were created to estimate the difference in risk of conversion to biomarker positivity by Aβ-PET, tau-PET, and plasma pTau-217 respectively for participants who were negative for each biomarker at baseline controlling for age, sex, years of education, *APOE* ε4 carrier status, and parental history of AD. Coefficients from these models were exponentiated to provide Hazard Ratios (HR) detailing the difference in risk of conversion by eCRF group.

LME models were used to evaluate the modification of the association between time-varying biomarker levels (per SD) and cognitive trajectory (PACC-3 score) by eCRF. These models were stratified by Aβ and tau status at any time point (Aβ-T-, Aβ+T-, Aβ-T+, and Aβ+T+) due to the potential synergistic effects of Aβ and T on cognitive change.[27,34–36] Models included baseline eCRF, biomarker level, years of follow-up, interaction terms between each of these variables, and a three-way interaction term between baseline eCRF, biomarker level, and years of follow-up. This three-way interaction term provided the coefficient of interest indicating differing rates of cognitive change per SD change in biomarker per year of follow-up based on eCRF group. These models were adjusted for baseline age, sex, years of education, APOE ε4 carrier status, parental history of AD, and random intercept and slope per participant.

## 3. Results

### 3.1 Participants

Baseline characteristics of the full sample and the Aβ+/- (by PiB-PET) sub-samples are provided in **Table 1**. The entire sample (N=533) was 70% female and majority of participants (79%) were Aβ-at baseline. These participants were younger, less likely to be *APOE* ε4 carriers or have a parental history of AD compared to those Aβ+ at baseline (all *p*s≤0.01; **Table 1**). Significant differences in baseline AD biomarker levels were observed between Aβ+ and Aβ-groups (all *p*s<0.001). The Aβ+ group also had significantly lower baseline PACC3 scores (*p*<0.001).

**Table 1:**
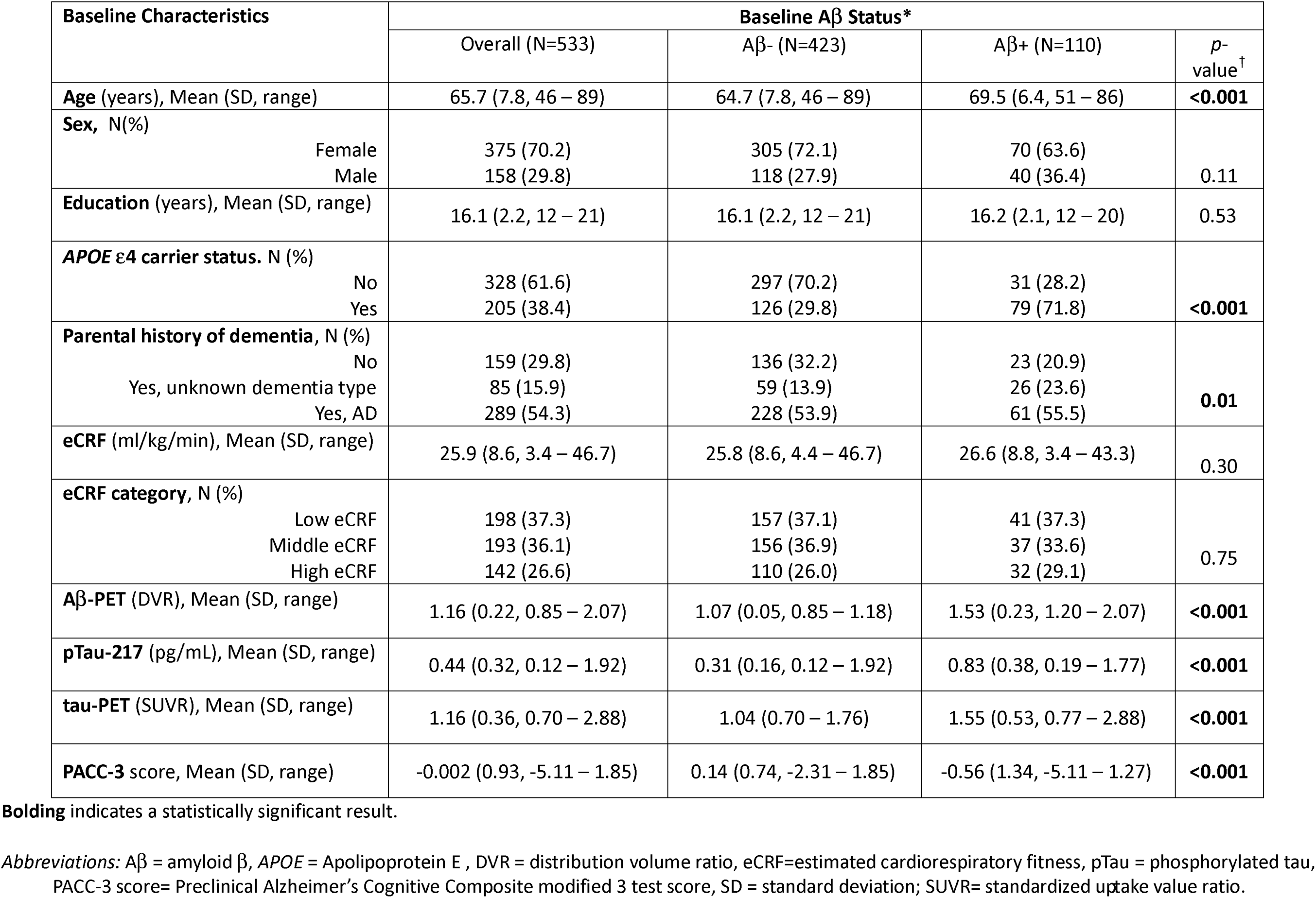
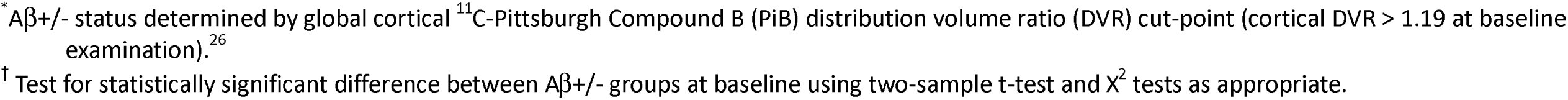
Baseline characteristics of the entire sample and by baseline Aβ status.

| Baseline Characteristics | Baseline Aβ Status* | | | |
| --- | --- | --- | --- | --- |
| | Overall (N=533) | Aβ- (N=423) | Aβ+ (N=110) | p-value<sup>†</sup> |
| **Age** (years), Mean (SD, range) | 65.7 (7.8, 46 – 89) | 64.7 (7.8, 46 – 89) | 69.5 (6.4, 51 – 86) | **<0.001** |
| **Sex**, N(%) |  |  |  |  |
| Female | 375 (70.2) | 305 (72.1) | 70 (63.6) | 0.11 |
| Male | 158 (29.8) | 118 (27.9) | 40 (36.4) |  |
| **Education** (years), Mean (SD, range) | 16.1 (2.2, 12 – 21) | 16.1 (2.2, 12 – 21) | 16.2 (2.1, 12 – 20) | 0.53 |
| **APOE ε4 carrier status**, N (%) |  |  |  |  |
| No | 328 (61.6) | 297 (70.2) | 31 (28.2) | **<0.001** |
| Yes | 205 (38.4) | 126 (29.8) | 79 (71.8) |  |
| **Parental history of dementia**, N (%) |  |  |  |  |
| No | 159 (29.8) | 136 (32.2) | 23 (20.9) | **0.01** |
| Yes, unknown dementia type | 85 (15.9) | 59 (13.9) | 26 (23.6) |  |
| Yes, AD | 289 (54.3) | 228 (53.9) | 61 (55.5) |  |
| **eCRF** (ml/kg/min), Mean (SD, range) | 25.9 (8.6, 3.4 – 46.7) | 25.8 (8.6, 4.4 – 46.7) | 26.6 (8.8, 3.4 – 43.3) | 0.30 |
| **eCRF category**, N (%) |  |  |  |  |
| Low eCRF | 198 (37.3) | 157 (37.1) | 41 (37.3) | 0.75 |
| Middle eCRF | 193 (36.1) | 156 (36.9) | 37 (33.6) |  |
| High eCRF | 142 (26.6) | 110 (26.0) | 32 (29.1) |  |
| **Aβ-PET** (DVR), Mean (SD, range) | 1.16 (0.22, 0.85 – 2.07) | 1.07 (0.05, 0.85 – 1.18) | 1.53 (0.23, 1.20 – 2.07) | **<0.001** |
| **pTau-217** (pg/mL), Mean (SD, range) | 0.44 (0.32, 0.12 – 1.92) | 0.31 (0.16, 0.12 – 1.92) | 0.83 (0.38, 0.19 – 1.77) | **<0.001** |
| **tau-PET** (SUVR), Mean (SD, range) | 1.16 (0.36, 0.70 – 2.88) | 1.04 (0.70 – 1.76) | 1.55 (0.53, 0.77 – 2.88) | **<0.001** |
| **PACC-3** score, Mean (SD, range) | -0.002 (0.93, -5.11 – 1.85) | 0.14 (0.74, -2.31 – 1.85) | -0.56 (1.34, -5.11 – 1.27) | **<0.001** |

**Bolding** indicates a statistically significant result.
*Abbreviations:* Aβ = amyloid $\beta$ , APOE = Apolipoprotein E, DVR = distribution volume ratio, eCRF=estimated cardiorespiratory fitness, pTau = phosphorylated tau, PACC-3 score= Preclinical Alzheimer's Cognitive Composite modified 3 test score, SD = standard deviation; SUVR= standardized uptake value ratio.
\* Aβ +/- status determined by global cortical <sup>11</sup>C-Pittsburgh Compound B (PiB) distribution volume ratio (DVR) cut-point (cortical DVR > 1.19 at baseline examination).<sup>26</sup>
† Test for statistically significant difference between Aβ +/- groups at baseline using two-sample t-test and $\chi^2$ tests as appropriate.

### 3.2 eCRF and biomarker trajectories

Aβ accumulation rates based on global cortical PiB-PET DVR scores (all *p*s≥0.08; **Table 2**) or by plasma pTau-217 (all *p*s≥0.25; **Table 2)** did not significantly differ by eCRF group regardless of baseline Aβ status. This lack of association between eCRF and pTau-217 remained when analyses were re-stratified by baseline pTau-217 status (pTau-217+/-; data not shown). There were no significant differences in accumulation rates of tau-PET by eCRF. (all *p*s≥0.15; **Table 2**). This lack of association between eCRF and tau remained unchanged when re-stratifying analyses by baseline T+/- status based on tau-PET (data not shown).

**Table 2:** Difference in rates of biomarker accumulation (95% CI) per eCRF standard deviation and by eCRF group, stratified by baseline Aβ status.

| | Aβ- | Aβ+ |
| --- | --- | --- |
| **Aβ-PET** |  |  |
| N | 423 | 110 |
| Per SD eCRF* | -.000 (-.000, .000) | .001 (-.000, .002) |
| By eCRF category<sup>†</sup> |  |  |
| Low | Reference Group |  |
| Middle | .000 (-.005, .006) | .006 (-.012, .024) |
| High | -.003 (-.009, .003) | .016 (-.001, .033) |
| **Plasma pTau-217** |  |  |
| N | 356 | 89 |
| Per SD eCRF* | .000 (-.000, .000) | -.002 (-.005, .002) |
| By eCRF category<sup>†</sup> |  |  |
| Low | Reference Group |  |
| Middle | .005 (-.003, .014) | -.012 (-.087, .063) |
| High | -.002 (-.011, .007) | -.036 (-.107, .035) |
| **tau-PET** |  |  |
| N | 362 | 95 |
| Per SD eCRF* | -.000 (-.001, .000) | -.002 (-.005, .001) |
| By eCRF category<sup>†</sup> |  |  |
| Low | Reference Group |  |
| Middle | -.000 (-.010, .009) | -.024 (-.100, .052) |
| High | -.007 (-.018, .003) | -.010 (-.083, .062) |

*Abbreviations:* Aβ = amyloid-β ; APOE =Apolipoprotein E, CI = confidence interval, eCRF= estimated cardiorespiratory fitness, PET= positron emission tomography, pTau = phosphorylated tau, SD = standard deviation.
\* Linear mixed effects models adjusted for baseline age, sex, years of education, APOE ε4 carrier status, parental history of AD, years of follow-up, and random intercept and slope per participant. Estimates represent difference in rate of biomarker accumulation based on interaction term between baseline eCRF (per SD) and years of follow-up.
<sup>†</sup> Linear mixed effects models adjusted for baseline age, sex, years of education, APOE ε4 carrier status, parental history of AD, years of follow-up, and random intercept and slope per participant. Estimates represent difference in rate of biomarker accumulation based on interaction term between baseline eCRF category and years of follow-up.

### 3.3 eCRF and biomarker conversion

For those Aβ-at baseline, the high eCRF group had a lower risk of conversion to Aβ+ status over the course of follow-up, with only 5.8% of those in the high compared to 10.8% in the low eCRF group converting. A Kaplan-Meier survival analysis confirmed a significantly lesser risk of conversion to Aβ+ status over the course of follow up for the high (*p*<0.001), but not the middle (*p*=0.18), eCRF group; **Figure 1**. Cox-proportional hazards model, adjusted for age, sex, years of education, *APOE* e4 carrier status, and parental history of AD, yielded similar results, whereby the high eCRF group had lesser risk (HR [95% CI = 0.42 [0.20, 0.88]) of conversion to Aβ+ compared to low eCRF group, indicating a 58% decrease in risk of conversion. No significant differences in risk were observed for the middle compared to the low eCRF group (HR [95%CI] = 0.85 [0.46, 1.54]; **Table 3**). Among those who were pTau-217-at baseline, the high eCRF group had a significantly lower risk of conversion via both Kaplan-Meier survival analysis (*p*=0.04) and Cox proportional hazards model (HR[95%CI]= 0.45 [0.20, 0.97]), which revealed a 55% risk reduction; **Figure 1, Table. 3** For those who were T-at baseline, Kaplan-Meier survival analysis indicated a trend toward better survival (non-conversion to T+); for both the middle and the high eCRF groups (all *ps* = 0.07); **Figure 1**. This association was not confirmed, however, by multivariable adjusted Cox proportional hazards modeling (all *p*s≥0.11; **Table 3**).

**Figure 1:**
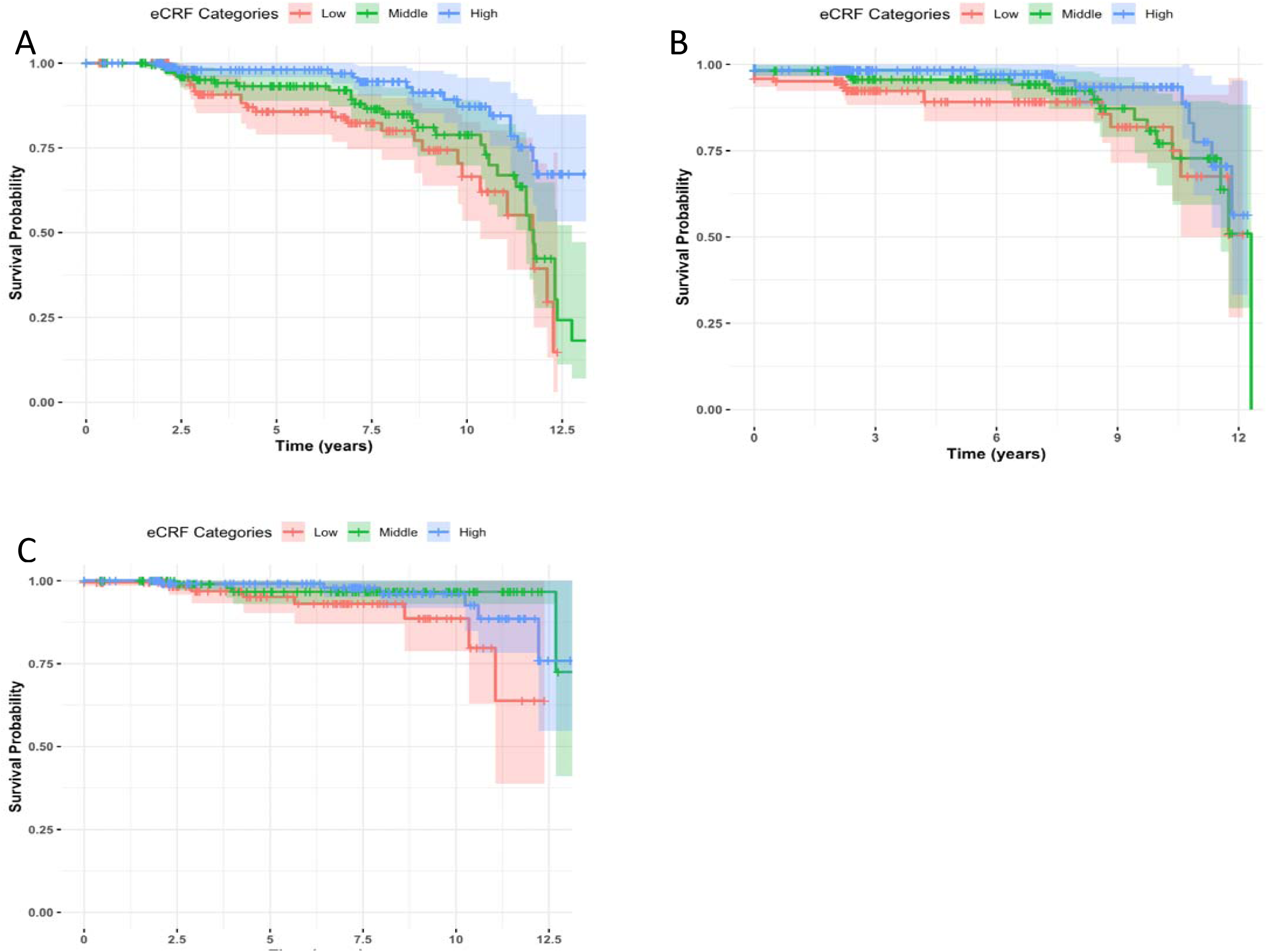
Kaplan-Meier curves depicting risk of becoming positive for core AD biomarkers over the course of follow-up among participants who were biomarker negative at baseline by eCRF group. Significantly lower risk of becoming positive for Aβ by PET (*p*=<0.001) and pTau-217 (*p*=0.04) was observed for high (but not the middle) compared to low eCRF group. A trend for lower risk of conversion to T+ was observed for the middle and high eCRF groups compared to the low eCRF group, though these relationships were not statistically significant (all *ps*>0.05). A: Aβ+ defined by a cortical PET-PiB DVR > 1.19. B: pTau-217+ defined by plasma pTau-217> 0.63 pg/mL. C: T+ defined by an entorhinal PET-^18^F-MK6240 SUVR> 1.27 or regional tau positivity in NFT associated regions corresponding to Braak staging. *Abbreviations:* Aβ = amyloid-β, eCRF = estimated cardiorespiratory fitness, NFT = neurofibrillary tangles, PiB DVR = ^11^C Pittsburgh Compound B distribution volume ratio, PET= positron emission tomography, pTau = phosphorylated tau, SUVR = standard uptake value ratio.

**Table 3:** Risk of conversion to biomarker positive status during follow-up by eCRF category among participants who were biomarker negative at baseline.*, ^†^.

| Group | Risk of Conversion, HR (95% CI) |  |  |
| --- | --- | --- | --- |
|  | Aβ-PET | Plasma pTau-217 | tau-PET |
| Low eCRF | Reference Group |  |  |
| Middle eCRF | 0.85 (0.46, 1.15) | 0.61 (0.31, 1.17) | 0.32 (0.08, 1.32) |
| High eCRF | **0.42 (0.20, 0.88)** | **0.45 (0.21, 0.97)** | 0.38 (0.09, 1.56) |

**Bolding** indicates a statistically significant result.
*Abbreviations:* Aβ= amyloid-β; CI= Confidence Interval, HR= Hazard Ratio, eCRF= estimated cardiorespiratory fitness, PET= positron emission tomography, pTau= phosphorylated tau.
\* Aβ+ determined by cortical PiB-DVR>1.19<sup>26</sup>; pTau-217+ determined by pTau-217>0.63 pg/mL<sup>29</sup>; T+ determined by entorhinal <sup>18</sup>F-MK6240 SUVR > 1.27 or AD-like positive Braak staging in any region based on PET MK6240 uptake.<sup>5,27,28</sup>
† Based on Cox-proportional hazards models adjusted for baseline age, sex, years of education, APOE ε4 carrier status, parental history of AD, baseline eCRF. Estimates represent risk of conversion to biomarker positivity.

### 3.4 eCRF, biomarker trajectories and cognitive change

In analyses stratified by Aβ/T status, we found a positive association between eCRF and cognitive trajectories among those who were Aβ+/T+, where high eCRF attenuated the negative association between higher levels of Aβ (per SD increase) and steeper cognitive decline; **Table 4**, **Figure 2**. Using estimates from the full model to calculate rates of cognitive change by eCRF holding covariates constant, the low eCRF group among Aβ+T+ had a decline in cognitive performance of -0.132/year per SD increase in Aβ, whereas those in the middle and high eCRF groups had significantly slower rates of cognitive decline (-0.023/year and -0.013/year per SD increase in Aβ, respectively, all *p*s*<0.05*). No significant relationships were observed between eCRF and cognitive decline for Aβ-T-, Aβ+T-, or Aβ-T+ groups (all *p*s≥0.24).

**Figure 2:**
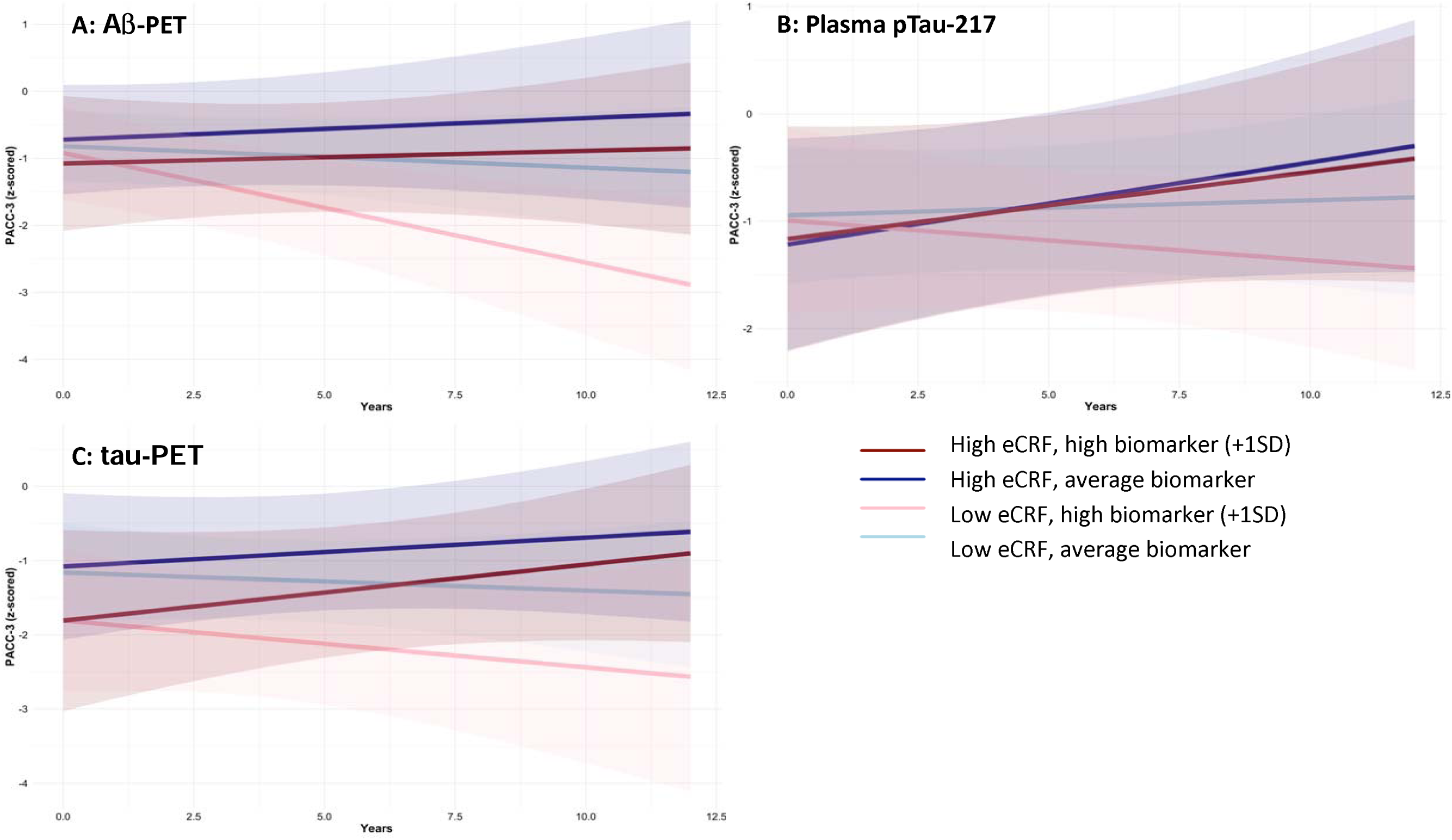
Differences in rates of cognitive change (PACC3 score) in Aβ+/T+, by eCRF category and core AD biomarker levels. The figures display PACC3 trajectory for the low (faint lines) and high (bolded lines) eCRF groups with different levels of each biomarker (average and 1SD above average). A statistically significant interaction was observed for Aβ-PET (Panel A; *p*<0.05); those with both high eCRF and high Aβ (A; dark red line) experienced no decline in cognition, whereas those with low eCRF and high Aβ (panel A; pink line) experienced significant decline. Similar trends were observed for plasma pTau-217 (B; *p*=0.31) and tau-PET (C; *p*=0.27,), though these results were not statistically significant. Aβ+/- status defined by a cortical PET-PiB DVR > 1.19, T+/- status defined by entorhinal PET-18F-MK6240 SUVR>1.27 or regional tau positivity in NFT associated regions corresponding to Braak staging. *Abbreviations:* Aβ = amyloid-β, eCRF = estimated cardiorespiratory fitness, NFT = neurofibrillary tangles, PET = positron emission tomography, PACC3 = Preclinical Alzheimer’s Cognitive Composite modified 3 test score, PiB DVR = 11CPittsburgh Compound B distribution volume ratio, SD = standard deviation, SUVR =standardized uptake value ratio. Figures were created using the coefficients from multivariable linear mixed effects models, adjusted for baseline age, sex, years of education, APOE ε4 carrier status, parental history of AD, baseline eCRF, years of follow-up, random intercept, and random slope per participant.

**Table 4:** Differences in rates of cognitive decline (PACC-3) by eCRF category, stratified by Aβ/T status.*, ^†^.

| | Aβ-T- | Aβ+T- | Aβ-T+ | Aβ+T+ |
| --- | --- | --- | --- | --- |
| Cognitive rate of change per SD increase in Aβ-PET<br>(difference in rate of change per year) | N=323 | N=64 | N=39 | N=85 |
| Low eCRF | Reference Group |  |  |  |
| Middle eCRF | -.005 (-.042, .032) | -.141 (-.374, .093) | .072 (-.180, .325) | **.109 (0.001, .217)** |
| High eCRF | -.000 (-.036, .036) | -.083 (-.316, .149) | .038 (-.092, .168) | **.119 (.020, .217)** |
| Cognitive rate of change per SD increase in pTau-217<br>(difference in rate of change per year) | N=263 | N=55 | N=36 | N=68 |
| Low eCRF | Reference Group |  |  |  |
| Middle eCRF | -.006 (-.046, .033) | -.119 (-.310, .072) | -.091 (-.507, .326) | 0.070 (-.030, .169) |
| High eCRF | -.004 (-.047, .040) | -.073 (-.266, .121) | **-.330 (-.473, -.186)** | 0.037 (-.034, 0.108) |
| Cognitive rate of change per SD increase in tau-PET<br>(difference in rate of change per year) | N=265 | N=49 | N=37 | N=80 |
| Low eCRF | Reference Group |  |  |  |
| Middle eCRF | -.042 (-.104, .020) | .015 (-.102, .133) | -.252 (-.683, .179) | .083 (-.119, .285) |
| High eCRF | -.003 (-.062, .057) | .038 (-.085, .161) | -.079 (-.476, .317) | .075 (-.059, .210) |

**Bolding** indicates a statistically significant result.
*Abbreviations:* Aβ = amyloid-β , AD= Alzheimer's disease, APOE = Apolipoprotein E, eCRF = estimated cardiorespiratory fitness, pTau = phosphorylated tau, PACC-3 score= Preclinical Alzheimer's Cognitive Composite modified 3 test score, PiB DVR= <sup>11</sup>C Pittsburgh Compound B distribution volume ratio, SUVR= standardized uptake value ratio.
\*Aβ + determined by cortical PET PiB DVR > 1.19; T+ determined by entorhinal <sup>18</sup>F-MK6240 SUVR > 1.27 or positive Braak staging in any region based on PET MK6240 uptake.
<sup>†</sup> Models adjusted for baseline age, sex, years of education, APOE ε4 carrier status, parental history of AD, baseline eCRF, years of follow-up, and random intercept per participant. Estimates represent difference in rate of change in PACC-3 score per SD increase in biomarker estimated by a three-way interaction term between baseline eCRF, years of follow-up, and z-scored biomarker level.

There were no significant associations between eCRF and cognitive decline with increasing plasma pTau-217 in Aβ-T- or Aβ+T-groups (all *p*s≥0.22). A significant negative association was observed in the Aβ-T+ group, whereby higher eCRF was associated with a steeper cognitive decline per SD increase in pTau-217 (*p*<0.01); **Table 4**. Among Aβ+T+, the middle and the high eCRF groups had slower cognitive decline compared to the low eCRF, though this association did not reach statistical significance (all *p*s≥0.17), **Table 4**, **Figure 2**.

No statistically significant relationships were observed between eCRF and cognitive decline with increasing levels of tau-PET (all *p*s≥0.19), albeit there was a non-significant trend for less cognitive decline with increasing tau burden in the middle and high eCRF groups among Aβ+T+ (all *p*s≥0.27), **Table 4**, **Figure 2**.

## 4. Discussion

In the current sample of middle-aged and older adults who were cognitively unimpaired at baseline from a cohort enriched for AD risk at enrollment, AD biomarker (Aβ and tau by PET, and plasma pTau-217) accumulation rates did not differ based on CRF. In those Aβ-at baseline, however, a 58% decrease in risk of conversion to Aβ+ status over the course of follow-up was observed for those with high compared to low CRF. Those with high CRF also had a 55% decrease in risk of conversion to pTau-217+. Importantly, the relationship between AD biomarker accumulation and cognitive decline seems to differ by CRF, such that for Aβ+T, high CRF was associated with slower rates of cognitive decline per SD increase in biomarkers.

The extant literature supports a protective effect of PA or CRF on brain health.[10,11,13,17,37] Higher levels of PA and higher CRF are associated with lower levels of core AD biomarkers,[11,12,38] although, some recent findings suggest no associations with biomarker deposition.[37,39,40] Our current results are in line with the more recent literature[37,39,40]– we too report no relationship between CRF and the rates of Aβ (by PET or plasma pTau-217) or tau (MK-6240 PET) accumulation. Of note, we stratified our sample based on baseline Aβ status, which no study reporting a significant association between PA or CRF and accumulation of core AD neuropathology previously accounted for. However, in those with high CRF who were Aβ-at baseline, we observed a ∼60% reduction in the risk of conversion to Aβ+ whether by PET or plasma pTau-217, both of which are indicative of clinically relevant Aβ accumulation. This is an important finding, as prevention or delay of accumulation to a point of clinical relevance could significantly reduce future negative sequalae of AD.

While CRF did not confer resilience against AD biomarker accumulation in our sample, we did see potential CRF contributions to resilience against cognitive decline. The relationship between AD biomarker accumulation and cognitive decline is well-established in the literature, whereby higher levels of AD biomarkers are associated with poorer cognitive performance[27,41] and precipitate cognitive decline.[30,34,35,42] Our current findings suggest that CRF may offset the deleterious effects of core AD biomarker accumulation on cognitive decline. Those who were Aβ+T+ but concomitantly had higher eCRF declined cognitively at a slower rate even when experiencing increasing levels of AD biomarkers. Although this finding was only statistically significant with regards to Aβ, similar associations were observed with pTau-217 and tau-PET accumulation and warrant further investigation. Conversely, our finding of an unexpected deleterious association between high eCRF and cognitive decline in the Aβ-T+ group with increasing pTau-217, could indicate that once a threshold of tau accumulation is crossed (T+), CRF may no longer confer resilience against cognitive decline, though due to the relatively small sample size in this group as well as the other subgroups (i.e., Aβ-T+ high eCRF), caution should be used in interpretation of this finding. Furthermore, while pTau-217 is considered a marker of brain Aβ, it is also indicative of other non-Aβ-related changes across the AD continuum, which might account for the disparity in the results between PiB-PET and plasma pTau-217.[29] Additionally, research has suggested that the cognitive trajectory of people with the Aβ-T+ biomarker profile, frequently termed suspected non-Alzheimer’s disease pathophysiology (SNAP),[43,44] follows more closely with those who are Aβ-T-than with those on the AD continuum,[45,46] though this relationship may be dependent on additional measures of neurodegeneration.[47] Our findings may reflect differences in the pathological processes that contribute to non-AD accumulation of tau, which may be robust to potential protective effects of CRF, or could be due to the unknown levels of other neurodegenerative markers (i.e., brain atrophy, neurofilament light chain),[43,47] that may be present in A-T+ but are outside the scope of the present study. This warrants further investigation with a larger sample of A-T+ individuals and with differentiation by additional neurodegenerative biomarkers.

While others have reported PA to be associated with better cognitive performance, they did not observe an attenuation of the adverse relationship between core AD pathology and cognition or did not evaluate the potential effect modification that CRF may have on this relationship.[14,24,48–50] Our current findings suggest that the deleterious relationship between accumulation of core AD neuropathology and cognitive decline could potentially be mitigated through exercise interventions and maintenance of CRF with age. In our sample, higher CRF seemed to be most beneficial for those participants who were Aβ+T+. Literature suggests that the “chronicity” of Aβ and tau positivity confers greater risk for and may have synergistic effects on cognitive decline.[5] Hence, the potential resilience conferred by CRF to those Aβ+T+ would be of utmost interest as a potential clinical intervention or preventative strategy.

One potential limitation of the current study is the use of an estimated measure of CRF, which relies on questionnaire data that could be prone to recall or reporting biases. Even so, this measure correlates well with VO_2peak_, the gold standard measure of CRF. [23,24] The WRAP and WADRC cohorts are predominantly highly educated and non-Hispanic white, making generalizability of the current findings unknown. Nonetheless, our findings are of value given the current lack of longitudinal studies on this and similar topics. The foregoing limitations notwithstanding, the WRAP and WADRC are prospective studies of well-characterized cohorts with high participation and retention rates and standardized procedures. This longitudinal design allows for characterization of biomarker trajectories and cognitive change, which is paramount to understanding the temporal relationships between potentially modifiable factors and AD-relevant outcomes.

### 4.1 Conclusions

Overall, our findings suggest that while CRF may not have an impact on the rate of accumulation of core AD biomarkers, high CRF does seem to reduce the risk of conversion from Aβ-to Aβ+. Perhaps more importantly, higher CRF seems to attenuate the otherwise deleterious relationship between core AD biomarker accumulation and cognitive decline, specifically for those at highest risk for AD (Aβ+T+). Together, our findings suggest an important role for CRF in relation to AD hallmark pathologic changes and offer a potential avenue for intervention.

## Data Availability

All data produced in the present study are available upon reasonable request to the authors, with center and institutional approval.

## Abbreviations

Aβ: amyloid-β;
AD: Alzheimer’s disease;
APOE ε4+: APOE ε4 allele carriage;
BMI: body mass index;
CRF: cardiorespiratory fitness
DVR: distribution volume ratio;
eCRF: estimated cardiorespiratory fitness,
HR: hazard ratio;
LME: linear mixed effects;
NFT: neurofibrillary tangle;
PA: physical activity;
PACC: Preclinical Alzheimer’s Cognitive Composite;
PET: positron emission tomography;
PiB: ^11^C-Pittsburgh Compound B;
pTau: phosphorylated tau;
RAVLT: Rey Auditory Verbal Learning Test Delayed Recall,
SD: standard deviation;
SNAP: suspected non-Alzheimer’s disease pathophysiology;
SUVR: standard uptake value ratios;
T: tau;
TMTB: Trails Making Test B;
VO_2peak_: peak oxygen consumption calculated for the present study using a validated eCRF equation;
WADRC: Wisconsin Alzheimer’s Disease Research Center;
WRAP: Wisconsin Registry for Alzheimer’s Prevention

## 5. Declarations of Interest

### Declarations of Interest

Dr. Ozioma Okonkwo receives grant support from the NIH. He also serves as the treasurer of the International Neuropsychological Society. He is also a guest editor of this journal but was not involved in the peer-review process of this article nor had access to any information regarding its peer-review. Drs. Ira Driscoll, Yue Ma, and Tobey Betthauser receive grant support from the NIH. Dr. Cynthia Carlsson receives grant support from the NIH, Department of Veterans Affairs, NIH/Lilly, NIH/Eisai, NIH/Cognition Therapeutics, and research medication support from Amarin Corporation. Dr. Kaj Blennow has served as a consultant, on advisory boards, or on data-monitoring committees for Acumen, ALZPath, AriBio, BioArctic, Biogen, Eisai, Lilly, Moleac Pte. Ltd, Novartis, Ono Pharma, Prothena, Roche Diagnostics, and Siemens Healthineers; has served on data monitoring committees for Julius Clinical and Novartis; has given lectures, produced educational materials, and participated in educational programs for AC Immune, Biogen, Celdara Medical, Eisai, and Roche Diagnostics; and is a co-founder of Brain Biomarker Solutions in Gothenburg AB, which is a part of the GU Ventures Incubator Program (outside submitted work). Dr. Henrik Zetterberg is a Wallenberg Scholar and a Distinguished Professor at the Swedish Research Council supported by grants from the Swedish Research Council (#2023-00356, #2022-01018 and #2019-02397), the European Union’s Horizon Europe research and innovation programme under grant agreement No 101053962, and Swedish State Support for Clinical Research (#ALFGBG-71320). Dr. Zetterberg has served at scientific advisory boards and/or as a consultant for Abbvie, Acumen, Alector, Alzinova, ALZpath, Amylyx, Annexon, Apellis, Artery Therapeutics, AZTherapies, Cognito Therapeutics, CogRx, Denali, Eisai, Enigma, LabCorp, Merry Life, Nervgen, Novo Nordisk, Optoceutics, Passage Bio, Pinteon Therapeutics, Prothena, Quanterix, Red Abbey Labs, reMYND, Roche, Samumed, Siemens Healthineers, Triplet Therapeutics, and Wave, has given lectures sponsored by Alzecure, BioArctic, Biogen, Cellectricon, Fujirebio, Lilly, Novo Nordisk, Roche, and WebMD, and is a co-founder of Brain Biomarker Solutions in Gothenburg AB (BBS), which is a part of the GU Ventures Incubator Program (outside submitted work). Dr. Sanjay Asthana receives grant support from the NIH. He also receives royalty as an editor of a textbook entitled, Hazzard’s Geriatrics and Gerontology; McGraw Hill, Publisher. Dr. Sterling Johnson receives grant support from the NIH. He also serves as a consultant and on advisory boards for ALZPath and Enigma Biosciences. All other authors have no relevant disclosures to report.

## 6. Consent Statement

Written informed consent was provided by all participants prior to study participation and approval was obtained from the institutional review board.

## 7. Funding

This research was supported by grants from the National Institute on Aging, R01AG062167 and R01AG085592 (Dr. Okonkwo), P30AG062715 (Dr. Asthana), RF1AG027161 (Dr. Johnson), R01AG054059 (Dr. Carey Gleason), and an NIH National Center for Advancing Translational Sciences grant UL1TR002373 awarded to the UW-Madison ICTR CTSA program. This study was supported in part by a core grant to the Waisman Center from the National Institute of Child Health and Human Development (P50 HD105353) and a NIH High-End Instrumentation grant (S10 OD030415). The funding sources for this work did not have a role in the study design, data analysis or interpretation, writing, or decision to submit this manuscript.

## 8. Author Contributions

AJP, BMB, and OCO designed the experiments. Data for the current project were provided by the laboratories of OCO, SCJ, SA, TJB, CLG, CMC, KB, and HZ. SRL and BMB collected and processed cardiorespiratory fitness data. ID, MPG, BTC and DBC were consulted on data acquisition, analyses or interpretation based on their respective expertise. AJP analyzed the data, wrote the manuscript, and prepared figures. All authors edited, revised, and approved the final version of the manuscript.

## 9. Acknowledgments

We would like to acknowledge and thank the staff and study participants of the Wisconsin Registry for Alzheimer’s Prevention and the Wisconsin Alzheimer’s Disease Research Center.

## Notes

### Author Declarations

The Health Sciences Institutional Review Board of the University of Wisconsin - Madison gave ethical approval for this work.

